# AI-Enabled Continuous Care Features in Real-World Psychotherapy: Treatment Engagement and Clinical Outcomes

**DOI:** 10.64898/2026.01.30.26345238

**Authors:** Scott Graupensperger, Millard Brown, Adam Chekroud, Baylee Mabe, Ondrej Kopecky, Nicole Srokosz, Jessica Hopkins, Matt Hawrilenko

## Abstract

**Importance:** AI-enabled features may improve the effectiveness of routine mental health care, yet large-scale real-world evidence remains limited.

**Objective:** To evaluate whether access to AI-enabled continuous care features embedded within routine psychotherapy delivery is associated with improved treatment engagement and clinical outcomes under real-world conditions.

**Design:** Preregistered cluster-level, matched, quasi-experimental study using a real-world rollout of AI-enabled continuous care features compared with psychotherapy alone (intention-to-treat framework).

**Setting:** An employer-sponsored behavioral health program providing outpatient psychotherapy for employees and dependents.

**Participants:** Adults initiating a new episode of psychotherapy from 25 employers with access to continuous care features and 75 matched employers without access. Treatment engagement was assessed over 7 weeks (*n*=26,208), and clinical outcomes were evaluated for up to 180 days (*n*=5,518).

**Exposure:** Employer-level access to AI-enabled continuous care features supporting engagement and continuity before and between psychotherapy sessions, compared with psychotherapy alone.

**Main Outcomes:** Early treatment engagement (number of psychotherapy sessions attended and time to second session) and changes in depressive and anxiety symptom severity measured using the Patient Health Questionnaire–9 (PHQ-9) and Generalized Anxiety Disorder–7 (GAD-7).

**Results:** Compared with matched controls receiving psychotherapy alone, the intervention group attended 5% more psychotherapy sessions during the first 7 weeks (rate ratio, 1.05 [1.01, 1.10]) and completed their second session sooner (mean difference, −0.62 days [−1.05, −0.18]). Both groups demonstrated substantial symptom improvement over time; however, access to continuous care features was associated with additional improvement in depressive symptoms (d=0.16) and anxiety symptoms (d=0.15) at the median duration of care (day 44). These effects translated into clinically meaningful differences in reliable improvement by the median duration of care (NNT=25 for both outcomes).

**Conclusions and Relevance:** In this real-world evaluation, access to AI-enabled continuous care features embedded within routine psychotherapy delivery was associated with greater early engagement and a higher likelihood of reliable symptom improvement beyond psychotherapy alone. These findings suggest that augmenting routine psychotherapy with AI-enabled continuous care can meaningfully shift recovery trajectories during a standard treatment episode, strengthening early treatment momentum and improving outcomes at scale.

**Key Points:** *Question:* Is access to AI-enabled continuous care features embedded within routine psychotherapy delivery associated with improved treatment engagement and clinical outcomes under real-world conditions?

*Findings:* In this cluster-level, matched, quasi-experimental study of adults receiving psychotherapy within an employer-sponsored behavioral health program, access to AI-enabled continuous care features was associated with significantly greater early treatment engagement and faster improvement in depressive and anxiety symptoms compared with psychotherapy alone.

*Meaning:* AI-enabled support features may incrementally enhance the delivery and effectiveness of established psychotherapies when implemented as complements to routine care at scale.

## Background

In routine outpatient settings, psychotherapy is typically delivered through discrete, session-based encounters with limited continuity between visits.^1–3^ This episodic structure can create gaps in engagement and therapeutic momentum, particularly early in care when dropout risk is highest.^4,5^ Blended-care and digital-assisted models have attempted to address these gaps by incorporating structured online modules, automated supports, and therapist-guided digital tools alongside traditional sessions, maintaining clinical outcomes while enhancing care delivery.^6–10^ However, most prior digital augmentations have relied on static, protocol-driven content, limiting their ability to respond dynamically to patients’ evolving needs or to provide continuous, personalized reinforcement between sessions. The emergence of generative artificial intelligence (AI) introduces new opportunities to extend episodic psychotherapy through dynamic, conversational tools designed to reinforce engagement and continuity throughout an episode of care, expanding between-session support beyond the constraints of module-based blended care models.^11–13^

Unlike standalone digital therapeutics or chatbot-based interventions, AI-enabled tools can be designed to augment clinician-delivered psychotherapy by supporting participants and providers before and between sessions.^14^ Opportunities to enhance care begin prior to the first visit, when structured onboarding may help clarify presenting concerns, prior treatment experiences, and therapeutic goals. In contrast to static intake forms, AI-guided onboarding can use adaptive, conversational dialogue to generate personalized follow-up questions and deliver contextually responsive, empathically framed replies in real time. By acknowledging and elaborating on participants’ expressed concerns, this approach is intended to facilitate reflection, clarify treatment goals, and shape early expectations about the purpose and potential value of care prior to the first session.^15^ Following sessions, AI-generated summaries and action-oriented takeaways may support recall, reflection, and follow-through on clinician-directed strategies, reinforcing continuity between visits. Together, these features may strengthen early therapeutic alignment, maintain treatment momentum throughout an episode of care, and ultimately propel greater clinical impact.

Despite growing enthusiasm for integrating AI into mental health care, empirical evidence from large-scale, routine practice settings remains sparse.^16,17^ In particular, it is unclear whether AI-enabled augmentation strengthens early engagement or meaningfully alters clinical trajectories when situated within standard psychotherapy. Addressing this gap will help determine whether AI integration represents a substantive advance in care delivery with the potential to transform psychotherapy from an episodic service into a more continuous, adaptive system capable of enhancing therapeutic impact at scale.

### Current Study

The current study leveraged a large-scale, real-world rollout of a first phase of AI-enabled continuous care features across 25 employers, with comparisons to participants from 75 matched employers that did not receive access during the study period. Continuous care refers to a novel care delivery model featuring AI-enabled tools designed to support engagement and continuity before and between psychotherapy sessions. We hypothesized that access to continuous care features would be associated with greater treatment engagement within the program and greater symptom improvement over time compared to access to psychotherapy alone. See preregistration at AsPredicted #237134: https://aspredicted.org/s7vr-6m7k.pdf).

## Method

### Intervention

The intervention was delivered within an employer-sponsored behavioral health program (Spring Health), which has been described previously and has demonstrated improvements in depression, anxiety, and workplace outcomes.^18–23^ The program provides access to a predefined number of prepaid psychotherapy and/or medication management sessions with licensed clinicians (6–18 sessions, depending on employer plan level), with additional care available through in-network health plan coverage.

All participants in both intervention and comparison employers initiated psychotherapy from therapists with master’s- or doctoral-level licenses through this base program. The intervention evaluated in the present study consisted of three additional optional AI-enabled continuous care features layered on top of standard psychotherapy services: guided intake, session summaries, and session takeaways. Guided intake supported structured onboarding prior to the initial psychotherapy session through an AI-assisted conversational interface that generated empathetic responses and personalized follow-up questions based on participants’ inputs. Session summaries and takeaways provided post-session summaries and action-oriented follow-up content derived directly from the recorded therapy sessions. In addition to summarizing clinician-directed guidance, session takeaways included personalized journaling prompts and, when applicable, recommended skills-based exercises mapped to themes identified in the session. Detailed descriptions and implementation considerations are provided in the Supplement.

### Study Design

This preregistered study used a cluster-level, matched, quasi-experimental design, with access to AI-enabled continuous care features implemented at the employer level rather than the individual level. Access to continuous care features was introduced through a planned, staged product rollout to 25 employers based on implementation timing and platform integration considerations. Because allocation was not randomized, these employers were matched to 75 employers without access using a 1:3 matching ratio. Matching was conducted at the employer level to improve comparability between intervention and comparison groups on key organizational characteristics, including size, industry, benefit design, and prior-year treatment-seeking rates; detailed matching procedures are provided in the Supplement.

Analyses were conducted using two prespecified analytic subsamples aligned with distinct study objectives. Treatment engagement was evaluated during a 7-week early engagement window following psychotherapy initiation, whereas clinical outcomes were evaluated over a longer follow-up period of up to 6 months. Observation windows and censoring rules were defined to ensure equal opportunity for follow-up and to minimize bias.

The study was preregistered prior to launch, and substantive deviations from the preregistered analysis plan and their implications for interpretation are described in the Supplement. This study involved secondary analysis of deidentified data generated from routine clinical care and program implementation, independent of research activities. Use of these data for research purposes is governed by an ongoing determination by the Yale Institutional Review Board that such analyses do not constitute human subjects research; accordingly, institutional review board approval and informed consent were not required for this study.

### Participants and Eligibility

Participants included adults actively enrolled in the behavioral health program who completed a baseline assessment prior to initiating psychotherapy. Enrollment occurred on a rolling basis as participants initiated a new episode of program-based care, defined as either a first encounter or the first encounter following a 6-month washout period. Treatment engagement analyses were restricted to participants with a complete 7-week observation window following psychotherapy initiation (n=26,208). Clinical outcome analyses focused on participants with at least one follow-up symptom assessment after baseline (n=5,676), who contribute within-person symptom change information; however, all participants were retained in mixed-effects models under an intention-to-treat framework, including those without follow-up assessments. Participant inclusion across analytic subsamples is summarized in the study flow diagram (Figure 1). Baseline characteristics and post-matching balance for each analytic subsample are presented in Table S1.

**Figure 1.**
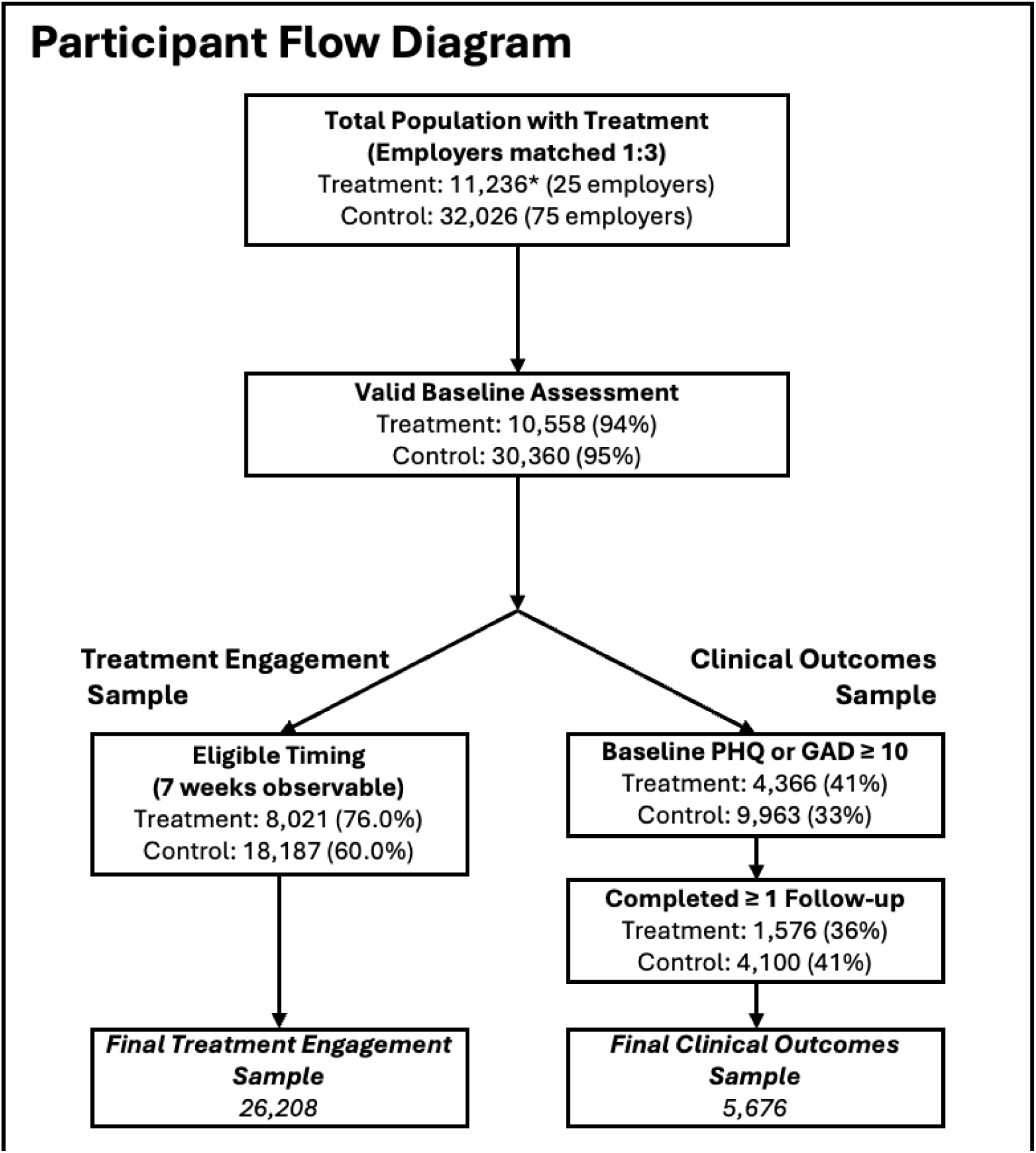
Eligible timing for treatment engagement models required 7 weeks of observable follow-up to avoid right-censoring. *39 treatment participants were removed from the sample because they were randomly selected to use an additional test-feature that was not part of the current study.

**Figure 2.**
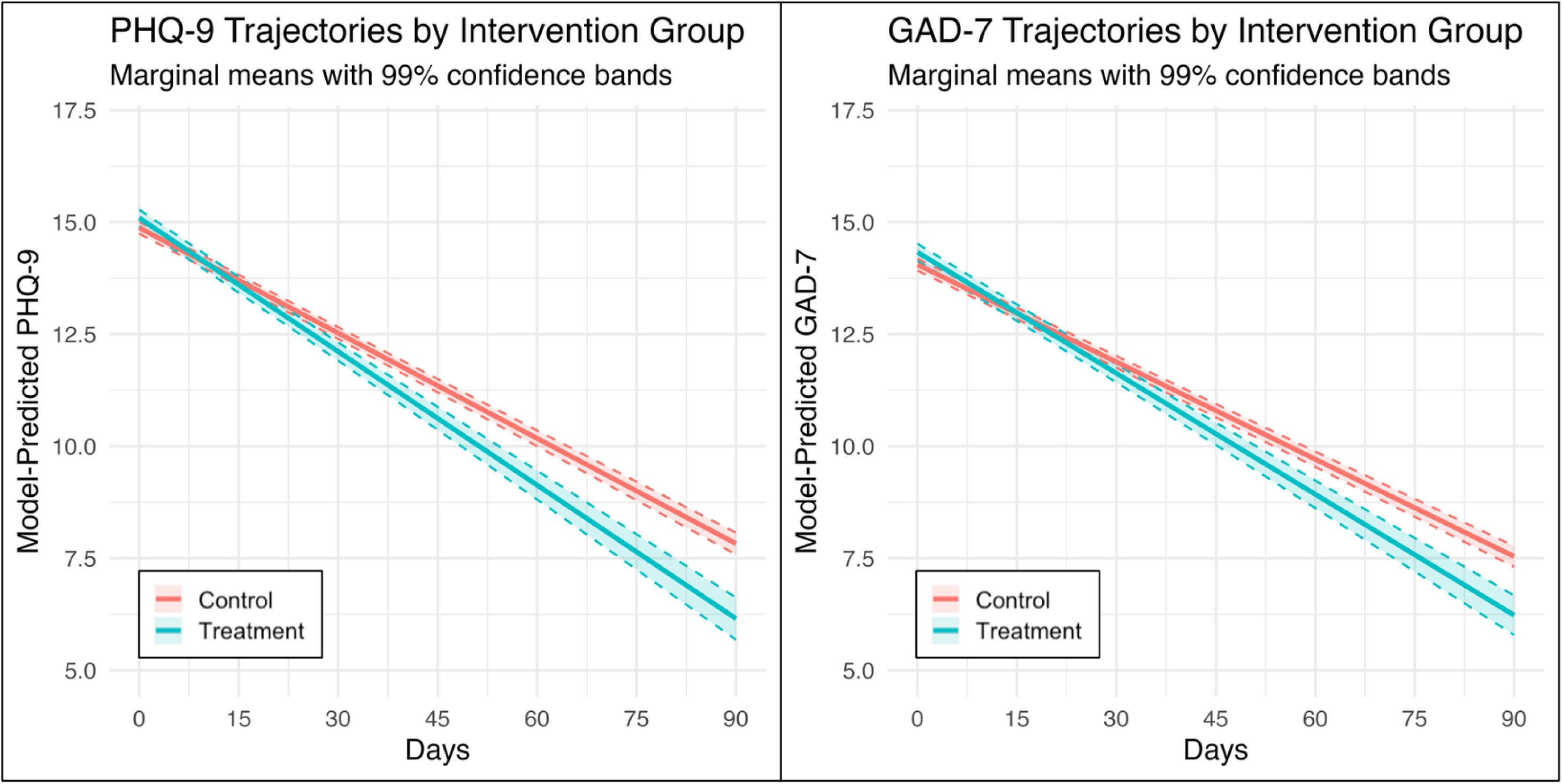
Model-predicted trajectories of depressive (PHQ-9) and anxiety (GAD-7) symptoms by intervention group. Lines represent marginal mean symptom scores estimated from mixed-effects linear models among participants with elevated baseline symptoms (PHQ-9 ≥1O or GAD-7≥10), adjusted for baseline severity, age, gender, and employer-level clustering. Shaded bands denote 99% confidence intervals. Differences in the rate of symptom improvement over time reflect the treatment x time interaction.

### Outcome Measures

Treatment engagement outcomes focused on early psychotherapy utilization and treatment momentum. Early utilization was defined as the number of attended psychotherapy sessions within 7 weeks of the initial session, and treatment momentum was assessed using time to second psychotherapy session among participants with at least two attended sessions (n=21,116; 81% of total sample).

Therapeutic alliance was assessed using an optional, session-level alliance survey administered via e-mail following psychotherapy visits. For participants with available data, therapeutic alliance was operationalized as the participant-level mean of all alliance scores collected within the 7-week study period. Response rates were low (10.3% of participants had at least one report, n=2,705).

Clinical outcomes included depressive and anxiety symptom severity, assessed using the Patient Health Questionnaire–9 (PHQ-9^24^) and the Generalized Anxiety Disorder–7 (GAD-7^25^), respectively. Symptom measures were collected at baseline and during routine measurement-based care follow-up. Additional details on outcome measures are provided in the Supplement.

### Statistical Analyses

Analyses were conducted under an intention-to-treat framework based on employer-level intervention assignment. Treatment engagement and clinical outcomes were compared between intervention and matched comparison groups using regression models appropriate for each outcome, with clustering accounted for at the employer level. Longitudinal clinical outcomes were analyzed using mixed-effects models including treatment, time, and their interaction. To contextualize clinical significance, we estimated the population-average probability of clinically reliable improvement at the median follow-up time (44 days), defined as a reduction of ≥5 points on the PHQ-9 or ≥4 points on the GAD-7 from baseline.^26,27^ Numbers needed to treat were calculated as the inverse of the absolute difference in improvement probabilities between groups. Sensitivity analyses included difference-in-differences models and falsification tests using a pre-implementation comparison cohort (n=36,294) drawn from the same 100 employers and meeting identical inclusion criteria in the year prior to rollout. Full analytic details are provided in the Supplement.

## Results

Baseline demographic and clinical characteristics were balanced between intervention and matched control groups across both analytic subsamples, with standardized mean differences below conventional thresholds for imbalance (Table S1). Baseline depressive and anxiety symptom severity was highly comparable between groups, with mean baseline PHQ-9 and GAD-7 scores differing by less than 0.1 points between intervention and comparison participants, supporting the comparability of cohorts prior to outcome assessment.

Among participants in intervention employers, utilization of continuous care features varied across individuals and features (Table S2). In the treatment engagement analytic sample, 38.2% of participants used at least one continuous care feature during the early engagement period, whereas uptake was higher in the clinical outcomes analytic sample (47.0%). Lower utilization was observed for features requiring additional provider and participant consent (Table S3). Exploratory analyses indicated that feature utilization was not strongly associated with baseline symptom severity or demographic characteristics (Table S4). Additional descriptive details, including provider- and participant-level opt-in rates for session recordings, are provided in the Supplement.

### Treatment Engagement Outcomes

In intent-to-treat analyses, intervention assignment was associated with statistically significant improvements in early therapy engagement (Model A in Table 1). Participants in the intervention group completed a higher number of therapy sessions during the first 7 weeks of care compared with controls (rate ratio = 1.05 [95% CI: 1.01, 1.10], *p*=.017), corresponding to a 5% relative increase in expected session counts after adjustment for baseline symptom severity, age, gender, and employer-level clustering. Model-implied mean session counts were 3.30 sessions for the intervention group and 3.13 sessions for the control group over the 7-week study period. The intervention group also had faster follow-up after treatment initiation. Among participants with at least two completed therapy sessions during the 7-week study period, those in the intervention group completed their second session an average of 0.62 days sooner than controls (β = −0.62 days [−1.05, −0.18], p=.006). Model-implied estimates indicated a mean time to second session of 12.39 days in the intervention group compared with 13.01 days in the control group.

**Table 1.** Early Treatment Engagement Outcomes During the First 7 Weeks of Care: Primary and Sensitivity Analyses.

| Model | Total Therapy Sessions<br>(7-week Study Period) |  |  | Time to Second Therapy Session<br>(Among those with 2+) |  |  | Therapeutic Alliance Mean Score<br>(7-week Study Period) |  |  |
| --- | --- | --- | --- | --- | --- | --- | --- | --- | --- |
|  | Rate Ratio | (95% CI) | p-value | β (days) | (95% CI) | p-value | β | (95% CI) | p-value |
| **A. Primary Intervention Models:** |  |  |  |  |  |  |  |  |  |
| Treatment (vs Control) | 1.05 | (1.01, 1.10) | .017 | -0.62 | (-1.05, -0.18) | .006 | -0.03 | (-0.22, 0.15) | .722 |
| *Sensitivity Analyses* |  |  |  |  |  |  |  |  |  |
| **B. Falsification Models:** |  |  |  |  |  |  |  |  |  |
| Treatment (vs Control) | 0.99 | (0.96, 1.03) | .688 | 0.16 | (-0.26, 0.57) | .466 | 0.03 | (-0.05, 0.11) | .430 |
| **C. Difference-in-Differences Models:** |  |  |  |  |  |  |  |  |  |
| Treatment × Phase Interaction (DiD estimate) | 1.05 | (1.01, 1.08) | .005 | -0.53 | (-0.97, -0.08) | .020 | -0.04 | (-0.17, 0.10) | .574 |

**Note:** Panel A presents primary intent-to-treat estimates of the association between treatment assignment and early therapy engagement outcomes. Panel B presents falsification (placebo) analyses applying the same model specifications to a pre-intervention period in which no treatment effect is expected. Panel C presents difference-in-differences sensitivity analyses estimating the treatment-by-period interaction. Total therapy sessions were modeled using Poisson mixed-effects regression, with rate ratios representing relative differences in the expected number of sessions during the 7-week study period (e.g., a rate ratio of 1.05 corresponds to a 5% relative increase). Time to second therapy session was modeled using linear mixed-effects regression among participants who completed at least two sessions during the study period. Therapeutic alliance was operationalized as the participant-level mean of all alliance survey scores collected within 49 days (approximately 7 weeks) following the first therapy session and was modeled using linear mixed-effects regression; coefficients represent differences in alliance score units. All models adjusted for baseline PHQ-9, baseline GAD-7, age, and gender, and included a random intercept for employer.

Therapeutic alliance was examined among the ∼10% of participants with available alliance survey data. Average alliance scores were high and similar between intervention and comparison groups (means 9.13 and 9.12 out of 10, respectively), and the intervention effect was non-significant (Table 1).

Sensitivity analyses leveraging a temporally shifted falsification cohort and difference-in-differences specifications yielded results consistent with the primary findings across all outcomes (Models B and C in Table 1). Together, these analyses provide convergent evidence that observed effects were attributable to the intervention rather than to secular trends or unobserved employer-level differences.

### Clinical Outcomes

Across both intervention and comparison groups, participants experienced substantial reductions in depressive and anxiety symptoms over the course of care, consistent with the expected benefits of psychotherapy. For reference, model-predicted PHQ-9 scores in the matched control group declined by 7.1 points, on average, by 90 days post-baseline, with a similar model-predicted reduction of 6.5 points on GAD-7 scores.

Within this context of effective base care, assignment to the continuous care features was associated with additional, incremental improvements in symptom trajectories over time. In primary intent-to-treat analyses among participants with elevated baseline symptoms (PHQ-9 ≥10 or GAD-7 ≥10) the group×time interactions indicated faster symptom reduction among intervention participants relative to matched-controls for both outcomes (Table 2; Figure 2).

**Table 2.** Clinical Symptom Trajectories Among Participants With Elevated Baseline Symptoms: Primary and Sensitivity Analyses.

| Model | Depressive Symptoms<br>(Baseline PHQ-9 ≥10) |  |  | Anxiety Symptoms<br>(Baseline GAD-7 ≥10) |  |  |
| --- | --- | --- | --- | --- | --- | --- |
|  | β (days) | (95% CI) | p-value | β (days) | (95% CI) | p-value |
| **A. Primary Intervention Models:** |  |  |  |  |  |  |
| Treatment × Day Interaction | −0.021 | (−0.026, −0.014) | <.001 | −0.018 | (−0.022, −0.013) | <.001 |
| Marginal Effect at Median Length of Care (Day 44) | −0.71 | (−0.92, −0.49) | <.001 | −0.50 | (−0.71, −0.29) | <.001 |
| *Sensitivity Analyses* |  |  |  |  |  |  |
| **B. Falsification Models:** |  |  |  |  |  |  |
| Treatment × Day Interaction | −0.001 | (−0.003, 0.001) | .099 | 0.000 | (−0.001, 0.001) | .910 |
| **C. Difference-in-Differences Models:** |  |  |  |  |  |  |
| Treatment × Day × Phase Interaction (DiD estimate) | −0.019 | (−0.025, −0.014) | <.001 | −0.018 | (−0.023, −0.013) | <.001 |

*Note:* Panel A presents primary intent-to-treat mixed-effects models estimating treatment-by-time interactions, with marginal treatment effects evaluated at the observed median length of care for these samples (44 days). Panel B presents falsification analyses applying identical model specifications to a pre-intervention period in which no treatment effect was expected. Panel C presents difference-in-differences sensitivity analyses estimating the treatment-by-time-by-period interaction. All models adjusted for baseline symptom severity, age, and gender, and included random intercepts for employer and participant.

For depressive symptoms, the intervention was associated with an additional reduction of 0.021 PHQ-9 points per day over controls. At the median duration of care (44 days), this corresponded to a marginal effect of −0.71 PHQ-9 points, reflecting a small standardized effect size (Cohen’s d=0.16). For anxiety symptoms, intervention was associated with an additional reduction of 0.018 GAD-7 points per day. At the median duration of care (44 days), the marginal effect was −0.50 GAD-7 points (d=0.15). To contextualize the clinical significance of these effects, we estimated the probability of reliable improvement at the median duration of care (44 days). The probability of reliable improvement on the PHQ-9 (≥5-point reduction) was 53.2% in the intervention group compared with 49.2% in matched controls (NNT=25). For GAD-7 (≥4-point reduction), the corresponding probabilities were 53.1% and 49.1% (NNT=25).

Falsification analyses applying identical trajectory models to a pre-intervention period showed no evidence of differential symptom improvement by intervention assignment for either outcome. In these placebo models, the group×time interaction was small and not statistically significant for both outcomes. Difference-in-differences sensitivity analyses further supported the primary findings, as intervention assignment was associated with a significantly greater rate of symptom improvement in the post-intervention period relative to the pre-intervention period for both depressive and anxiety symptoms. The group×time×phase interaction indicated an additional reduction of 0.019 PHQ-9 points per day (*p*<.001) and 0.018 GAD-7 points per day (*p*<.001) attributable to the intervention (See Figure S1).

### Baseline Severity as a Moderator of Intervention Effects

In models including three-way interactions between intervention assignment, time, and baseline symptom severity, higher baseline PHQ-9 and GAD-7 scores were associated with significantly stronger effects (both interaction estimates *p*<.001). Probing model-estimated trajectories indicated that intervention effects were driven by patients with higher baseline severity (Figure 3). For example, participants with a baseline score of 10 experienced comparable effects to the control group at the median length of care, whereas participants with a baseline score of 20 had 1.69 [−1.99, −1.40] point larger decreases in PHQ-9 (Cohen’s *d*=0.29) and 1.48 [−1.84, −1.13] point larger decreases in GAD-7 (*d*=0.30) relative to matched-controls. Together, these patterns indicate greater clinical benefit among participants with more severe baseline symptoms.

**Figure 3.**
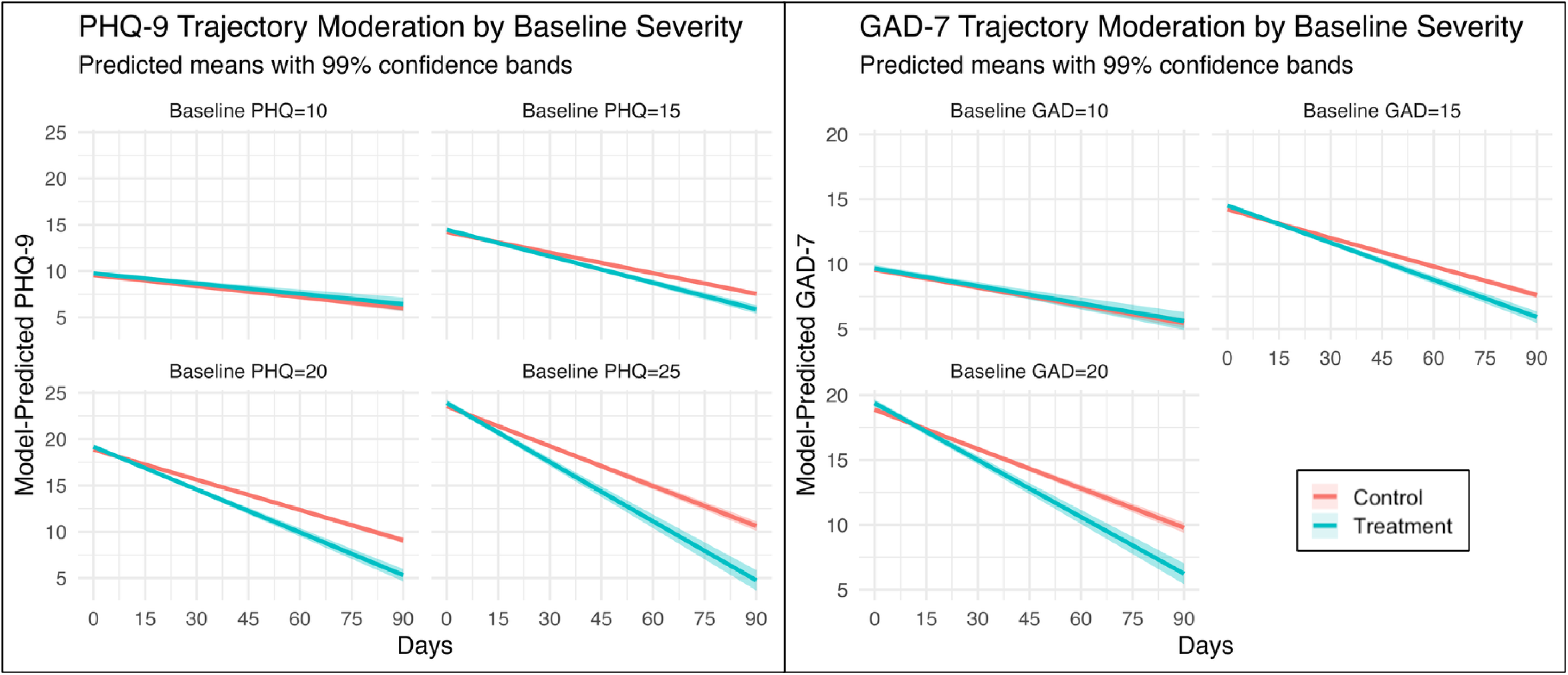
Model-predicted trajectories of depressive (PHQ-9) and anxiety (GAD-7) symptoms by baseline symptom severity. Lines represent marginal mean symptom scores estimated from mixed-effects linear models among participants with elevated baseline symptoms (PHQ-9 ≥ 10 or GAD-7 ≥ 10), adjusted for baseline severity, age, gender, and employer-levelclustering. Shaded bands denote 99% confidence intervals. Facets display predicted trajectories at representative baseline severity levels, illustrating greater intervention-associatedimprovement over time among participants with higher baseline symptom burden.

## Discussion

In this large-scale evaluation of AI-enabled continuous care features embedded within an employer-sponsored behavioral health program, access to these features integrated into psychotherapy was associated with improved treatment engagement and greater symptom improvement compared with psychotherapy alone. Under routine implementation conditions, these findings demonstrate that lightweight, AI-enabled supports can enhance psychotherapy delivery at scale, even when layered onto an already effective base of care. Although the incremental effects were modest in magnitude (*d*=0.15-0.16), this corresponded to approximately one additional reliable improvement for every 25 participants with access to continuous care over a median 44-day treatment episode.

Improvements in clinical outcomes were accompanied by meaningful differences in early treatment engagement, suggesting a plausible mechanism through which the continuous care features may exert their effects. The intervention group completed 5% more therapy sessions during the 7-week follow-up period and progressed more quickly from initial to follow-up visits, indicating denser and more rapid engagement during a critical treatment window, despite no significant impact on therapeutic alliance. These findings suggest that the features may operate through mechanisms other than therapeutic alliance. Guided intake may facilitate feelings of being heard, understood, and supported prior to the first session, while fostering positive treatment expectancy and readiness to engage in care.^28–30^ Between-session supports may operate through a distinct pathway by strengthening memory for treatment content and facilitating skill rehearsal. Such processes have been shown to predict symptom change independent of patients’ perceived bond with the therapist.^31–33^ It is also possible that, given alliance scores were uniformly near the ceiling across both groups, there was little opportunity to detect incremental differences.

Intervention effects were largest among individuals with higher baseline severity. This common pattern^34^ suggests that incremental support features integrated into psychotherapy may be most beneficial for individuals with greater initial clinical need. One potential explanation is that individuals with higher symptom severity may experience greater emotional activation during sessions and, as a result, have more difficulty retaining session content and benefit more from structured memory supportive interventions.^35,36^ Structured summaries and postsession takeaways may support consolidation and application of therapeutic material after the session has ended.

Taken together, these findings suggest that modest gains from lightweight, AI-enabled support features can yield meaningful clinical impact when deployed at scale. In contrast to psychotherapy enhancements that require substantial clinician training or sustained workflow changes, the continuous care features evaluated here can be implemented with minimal burden, such that even small incremental improvements may translate into meaningful population-level benefits.

## Limitations

We modified several elements of the preregistered analysis plan to reflect the scale and timing of the real-world rollout, including expanding the employer sample, adopting employer-level intent-to-treat analyses, and modeling symptom trajectories longitudinally rather than at a single post-treatment endpoint (see Table S5). These changes increased statistical power and strengthened causal interpretability while remaining consistent with the original study design and primary research questions. While the employer-level rollout and matched comparison design strengthen causal inference, the study was not randomized, and residual confounding cannot be fully excluded. Although employers were matched on prespecified organizational characteristics, residual confounding cannot be fully excluded. However, intervention and comparison employers exhibited similar pre-intervention outcome trends, and falsification and difference-in-differences sensitivity analyses yielded consistent results, reducing concern that selection into rollout explains the findings. Uptake of the AI-enabled features varied across participants, and the intent-to-treat framework does not permit isolation of effects attributable to specific features or levels of use when participants had access to all or no features. Therapeutic alliance was assessed via an optional, session-level survey with limited coverage, constraining inference regarding effects on relational processes. Finally, findings may not generalize beyond employees and dependents with access to employer-sponsored behavioral health benefits or to populations with severe mental illness or acute suicidal risk, who were not the focus of this evaluation.

## Conclusions

The present findings demonstrate that AI-enabled support features embedded within routine psychotherapy delivery can yield meaningful improvements in engagement and clinical outcomes under real-world conditions. Although these effects are incremental at the individual level, they are a lightweight addition and accrue on top of an already effective base of mental health care, translating into clinically meaningful population-level impact when deployed at scale. More broadly, these results showcase the value of evaluating digital augmentations as complements to clinician-delivered psychotherapy rather than as standalone interventions, and highlight the importance of rigorous, pragmatic evaluation as such tools continue to be integrated into mental health care delivery.

## Data Availability

The data used in this study include sensitive personal health information and are not publicly available. Deidentified data may be made available to qualified researchers upon reasonable request and subject to applicable legal, regulatory, and privacy requirements. Requests should be directed to the Yale Human Research Protection Program.

## Acknowledgements

**Disclosures:** All authors report being employed by and holding equity in Spring Health, Inc. outside the submitted work. In addition, Dr. Chekroud reported being the lead inventor on 3 patent submissions relating to treatment for major depressive disorder (US Patent and Trademark Office number Y0087.70116US00 and provisional application numbers 62/491 660 and 62/629 041) outside the submitted work. Finally, Dr. Chekroud reported holding equity in Carbon Health Technologies Inc, Wheel Health Inc, Parallel Technologies Inc, Healthie Inc, and UnitedHealthcare; and providing unpaid advisory services to health care technology startups outside the submitted work.

**AI-Assisted Technologies:** The authors used ChatGPT (OpenAI) during the research and writing process to assist with debugging statistical code and to suggest wording changes aimed at improving clarity and reducing word count. No data, analyses, figures, or images were generated by AI.

**Data Availability Statement:** The data used in this study include sensitive personal health information and are not publicly available. Deidentified data may be made available to qualified researchers upon reasonable request and subject to applicable legal, regulatory, and privacy requirements. Requests should be directed to the Yale Human Research Protection Program.

## Supplemental Materials

### Employer-Level Matching Procedures

Employers receiving access to continuous care features (“intervention employers”) were matched to employers without access (“comparison employers”) using a 1:3 matching ratio.

Matching was conducted at the employer level to reduce baseline differences between intervention and comparison groups and to improve comparability consistent with a cluster-randomized design, given that access to continuous care features was implemented at the employer rather than individual level.

Matching variables were prespecified and selected to capture key organizational characteristics plausibly associated with treatment access and engagement. These included number of covered lives (as a measure of employer size), industry, plan level (defined by the number of prepaid psychotherapy sessions included in the benefit), and prior treatment-seeking rate, operationalized as the proportion of covered lives with at least one behavioral health treatment encounter during the year preceding the study period. Matching was performed prior to participant-level outcome analyses and without reference to post-intervention outcomes.

### Rollout Timing and Censoring

Access to AI-enabled continuous care features was implemented in a staged employer-level rollout. Six intervention employers received access on July 8, 2025, followed by an additional 19 intervention employers on August 13, 2025. General availability of continuous care features to all employer customers, including comparison employers, occurred on December 3, 2025. To avoid contamination of the comparison condition, all analyses were censored at the date of general availability. That is, to ensure comparability between intervention and matched control groups, data collected after December 3rd, 2025 were not used in the present study.

### Outcome Definitions and Measurement

#### Early treatment engagement

Early treatment engagement outcomes were designed to capture initial uptake and momentum following psychotherapy initiation. Utilization during the early engagement window was operationalized as the number of attended psychotherapy sessions within 7 weeks of the initial session. Treatment momentum was additionally assessed using time to second psychotherapy session among participants with at least two attended sessions, as shorter time to follow-up reflects more rapid progression beyond initial access and is commonly used as a behavioral indicator of early engagement.

#### Therapeutic alliance

Therapeutic alliance was measured using an optional, session-level survey administered via email following psychotherapy visits. The survey was adapted from the Working Alliance Inventory and included three items reflecting core alliance domains: (1) bond (e.g., “I have a good working relationship with my provider”), (2) agreement on goals (e.g., “I understand and agree with the goals my provider and I have set”), and (3) agreement on tasks (e.g., “My provider and I agree on the best approach for addressing my problems”). Each item was rated on a 1–10 scale, with anchors of 1 (“not at all true”), 5 (“undecided”), and 10 (“completely true”). For participants with available data, alliance was operationalized as the participant-level mean of all alliance scores collected within the 7-week study period.

#### Clinical symptoms and reliable improvement

Depressive and anxiety symptom severity were assessed using the Patient Health Questionnaire–9 (PHQ-9) and the Generalized Anxiety Disorder–7 (GAD-7), respectively, collected at baseline and during routine measurement-based care follow-up. In addition to modeling continuous symptom trajectories, clinically reliable improvement was defined as a reduction of ≥5 points on the PHQ-9 or ≥4 points on the GAD-7 from baseline, consistent with prior validation studies. These thresholds were used to estimate the probability of reliable improvement at the median follow-up time in model-based analyses.

### Assessment of Post-Matching Balance

Following employer-level matching, balance between intervention and comparison groups was assessed at the participant level within each analytic subsample using standardized mean differences (SMDs). Balance was evaluated separately for the treatment engagement analytic sample (participants with a complete ≥49-day observation window) and the clinical outcomes analytic sample (participants with PHQ-9 or GAD-7 score ≥10 and at least one follow-up assessment), as these samples differed slightly in eligibility criteria and follow-up structure.

Table S1 presents baseline demographic and clinical characteristics for treatment and control participants within each analytic subsample. Across both subsamples, standardized mean differences for all assessed covariates were small in magnitude and below the conventional threshold of |SMD| < 0.10, indicating good balance between groups after matching.

**Table S1.** Baseline balance by analytic subsample after employer-level matching.

| **A. Treatment Engagement Analytic Sample (n = 26,208)** |  |  |  |
| --- | --- | --- | --- |
| **Characteristic** | **Treatment** | **Control** | **SMD** |
| Baseline PHQ-9, mean (SD) | 9.74 (6.3) | 9.75 (6.2) | -0.002 |
| Baseline GAD-7, mean (SD) | 9.31 (5.6) | 9.30 (5.5) | 0.003 |
| Age, mean (SD) | 37.40 (11.7) | 36.6 (11.7) | 0.066 |
| Man, % | 28.3% | 30.0% | -0.037 |
| Woman, % | 55.1% | 50.8% | 0.086 |
| Other / unknown gender, % | 16.6% | 19.2% | -0.067 |
| Dependent enrollee, % | 21.9% | 21.0% | 0.021 |
| **B. Clinical Outcomes Analytic Sample (n = 5,676)** |  |  |  |
| **Characteristic** | **Treatment** | **Control** | **SMD** |
| Baseline PHQ-9, mean (SD) | 13.85 (5.6) | 13.90 (5.7) | -0.008 |
| Baseline GAD-7, mean (SD) | 12.90 (4.7) | 13.01 (4.7) | -0.022 |
| Age, mean (SD) | 37.1 (12.0) | 36.5 (11.8) | 0.052 |
| Man, % | 27.4% | 27.8% | -0.009 |
| Woman, % | 56.2% | 53.8% | 0.049 |
| Other / unknown gender, % | 16.3% | 18.4% | -0.054 |
| Dependent enrollee, % | 21.4% | 22.0% | -0.014 |

**Note:** Values shown are standardized mean differences (SMDs), calculated as the difference in means or proportions between groups divided by the pooled standard deviation. Absolute SMD values less than 0.10 are commonly considered indicative of adequate balance between groups. To further account for any residual imbalance, all primary regression models are additionally adjusted for baseline covariates, including age, baseline symptom severity, and gender.

Across both analytic subsamples, baseline clinical symptom severity was highly comparable between treatment and comparison groups. SMDs for baseline PHQ-9 and GAD-7 scores were near zero in both the treatment engagement and clinical outcomes samples, indicating nearly identical baseline symptom severity across groups. Given that depressive and anxiety symptom severity represent the primary study outcomes and are most plausibly related to treatment engagement and improvement, this close balance provides particularly strong evidence of comparability between groups following employer-level matching.

Small residual differences were observed for select demographic characteristics, most notably gender composition, with standardized mean differences for women approaching but remaining below conventional thresholds for imbalance. All other covariates demonstrated standardized mean differences well below 0.10.

To further mitigate the potential influence of any residual baseline differences, all primary regression models were additionally adjusted for baseline covariates, including age, baseline symptom severity, and gender. This analytic approach combines employer-level matching with covariate adjustment at the participant level and is consistent with recommended best practices for observational and quasi-experimental studies. As a result, study findings reflect differences between groups that are robust to minor residual imbalances observed after matching.

### Detailed Description of Continuous Care Features and Session Recordings

Continuous care comprised three optional AI-enabled features designed to support engagement and continuity of care between psychotherapy sessions: guided intake, session summaries, and session takeaways. Each feature leveraged large language models to structure unstructured information, personalize content to the individual member, and reduce cognitive and logistical burden outside of live sessions.

**Guided intake** provided an AI-assisted conversational experience prior to the first psychotherapy session. Using a natural-language dialogue, the system prompted participants to describe their primary concerns, relevant history, symptoms, and treatment goals in their own words. AI models synthesized these free-text responses into a structured intake summary, organizing information into clinically relevant categories (e.g., presenting problems, goals, contextual factors). With participant consent, the intake summary was made available to the assigned provider ahead of the first session. The intended purpose of guided intake was to help participants reflect on and articulate their needs before initiating care, while also enabling providers to begin treatment with clearer context, potentially reducing time spent on administrative intake and supporting earlier therapeutic alignment.

Guided intake was introduced approximately one month into the study period (mid-August), such that participants initiating care early in the rollout period did not have the opportunity to use this feature prior to their first psychotherapy session, though it remained available thereafter. This staggered availability reflects real-world implementation conditions for digital care tools and, under the intent-to-treat analytic framework used here, would be expected to attenuate rather than inflate observed treatment effects.

**Session summaries** used AI to generate structured summaries of psychotherapy sessions that were recorded and transcribed with explicit consent from both participant and provider. Large language models processed session transcripts to identify key discussion themes, therapeutic focus areas, and high-level progress indicators, producing a concise, non-verbatim summary of each session. Summaries were delivered to participants after sessions to support recall, reflection, and continuity across appointments. By externalizing session content into a persistent artifact, the feature aimed to reinforce therapeutic insights, reduce reliance on memory alone, and help participants maintain momentum between sessions without requiring additional effort from providers.

**Session takeaways** complemented session summaries by generating brief, action-oriented follow-up points drawn directly from guidance, exercises, or recommendations articulated by the provider during the session. AI models identified these clinician-directed elements within the session transcript and translated them into clear, participant-facing takeaways, without introducing new clinical content. Takeaways were intended to support between-session engagement by clarifying provider-recommended next steps and encouraging application of therapeutic strategies discussed during the session. Whereas summaries emphasized reflection and continuity, takeaways emphasized behavioral follow-through and practical application of provider guidance.

All features were available to participants in intervention employers; however, feature use was optional and participant-driven. Uptake of continuous care features varied across participants and was lower than anticipated at the time of preregistration, particularly for session summaries and takeaways, which required additional consent and workflow integration. As a result, all primary analyses were conducted using an intent-to-treat framework based on employer-level access to continuous care features. Observed uptake rates by feature are summarized in Table S2.

**Table S2.** Utilization rates of the continuous care features.

| Continuous Care Feature | Treatment Engagement Sample | Clinical Outcomes Sample |
| --- | --- | --- |
| *Any continuous care feature use* | 38.2% | 47.0% |
| Used takeaways | 25.0% | 36.8% |
| Used session summaries | 15.6% | 22.7% |
| Used guided intake | 16.4% | 14.1% |

Access to session summaries and takeaways required that both the provider and the participant consent to having the psychotherapy session recorded and transcribed, with all identifying information removed prior to processing. Lower-than-anticipated opt-in rates—particularly among providers—limited the number of sessions eligible for these features and contributed to the observed utilization rates. Provider and participant opt-in rates are shown in Table S3.

**Table S3.** Provider and participant opt-in rates to session recordings (needed for transcript generation).

| Analytic Sample | Opt-In Metric | Provider Opt-In | Participant Opt-In |
| --- | --- | --- | --- |
| Treatment Engagement Sample | Session-level opt-in rate | 60.3% | 71.6% |
| Treatment Engagement Sample | Ever opt-in ($\geq 1$ session) | 67.3% | 85.6% |
| Treatment Engagement Sample | First-session opt-in rate | 56.9% | 74.7% |
| Clinical Outcomes Sample | Session-level opt-in rate | 59.9% | 73.8% |
| Clinical Outcomes Sample | Ever opt-in ($\geq 1$ session) | 67.2% | 89.8% |
| Clinical Outcomes Sample | First-session opt-in rate | 55.6% | 76.2% |

### Statistical Analyses

All analyses were conducted at the participant level under an intention-to-treat framework based on employer-level assignment to AI-enabled continuous care features. All regression models adjusted for baseline symptom severity, age, and self-reported gender. Treatment engagement outcomes were analyzed using regression models appropriate for the distribution of each outcome (e.g., negative binomial models for count outcomes). Employer-level clustering was accounted for using random intercepts.

Clinical outcomes were analyzed using mixed-effects regression models to account for repeated symptom assessments within participants and clustering within employers. Models included fixed effects for treatment group, time since psychotherapy initiation, and their interaction, with random intercepts for both participants and employers. Participants without follow-up assessments were retained in all models to preserve the intention-to-treat estimand, but only those with follow-up data contributed to the symptom trajectory coefficients of interest.

To estimate clinically interpretable effects, population-average predicted symptom trajectories were used to calculate the probability of clinically reliable improvement at the median follow-up time (44 days). Reliable improvement was defined as a reduction of ≥5 points on the PHQ-9 or ≥4 points on the GAD-7 from baseline. Individual-level probabilities were derived from model-implied outcome distributions using population-average predictions and averaged within treatment and comparison groups. Numbers needed to treat were calculated as the inverse of the absolute difference in improvement probabilities.

### Sensitivity Analyses

To evaluate the robustness of estimated treatment effects and assess the potential influence of residual confounding or secular trends, we conducted a series of prespecified sensitivity analyses designed to test key identifying assumptions of the primary analytic approach.

First, to further assess whether observed treatment effects could be attributed to time-varying confounding or cohort effects unrelated to the intervention, temporal falsification analyses were conducted using data from the same 100 employers during the year preceding intervention rollout. Identical model specifications—including covariates, random effects structure, and outcome definitions—were applied to this pre-implementation cohort (n=36,294), with a pseudo-implementation date assigned to mirror the timing of the actual rollout. Under this falsification test, detection of a treatment-associated effect in the pre-implementation period would suggest residual confounding, whereas null effects would provide evidence that post-implementation differences are unlikely to reflect secular trends or unobserved employer-level differences.

Difference-in-differences (DiD) models were estimated to compare changes in symptom trajectories between intervention and comparison groups before and after implementation of AI-enabled continuous care features. These models contrasted the effect found among the present intervention sample to a pre-implementation sample (i.e., the year prior to launch), where the DiD treatment effect was defined as the differential change in slopes following rollout. Under this framework, a null or comparable change in slopes between groups during the pre-implementation period would support the parallel trends assumption, whereas emergence of differential improvement only after implementation would be consistent with a causal intervention effect.

Together, these sensitivity analyses provide complementary tests of the core identifying assumptions underlying the primary analyses and support the interpretation that observed post-implementation differences in clinical outcomes reflect the effect of access to AI-enabled continuous care features rather than artifacts of timing, regression to the mean, or unmeasured baseline differences. All analyses were conducted using R (version 4.4.3).

### Predictors of Continuous Care Feature Utilization

To examine whether baseline clinical or demographic characteristics were associated with use of continuous care features among participants with access, we conducted an exploratory analysis restricted to participants in intervention employers. Any continuous care feature use was defined as use of at least one AI-enabled feature (guided intake, session summaries, or session takeaways) during the 7-week observation window. Associations between baseline characteristics and feature utilization were estimated using a mixed-effects logistic regression model with a random intercept for employer to account for clustering. Fixed-effect covariates included baseline PHQ-9 and GAD-7 scores, age, gender, and dependent enrollee status.

In this analysis of 8,021 participants nested within 25 employers, no strong associations were observed between baseline clinical severity or demographic characteristics and use of continuous care features (Table S4). Baseline PHQ-9 and GAD-7 scores were not associated with feature uptake, and effect estimates for age, gender, and dependent enrollee status were small in magnitude. Overall, these findings indicate that utilization of continuous care features among participants with access was not strongly patterned by baseline symptom severity or demographic factors.

**Table S4.**
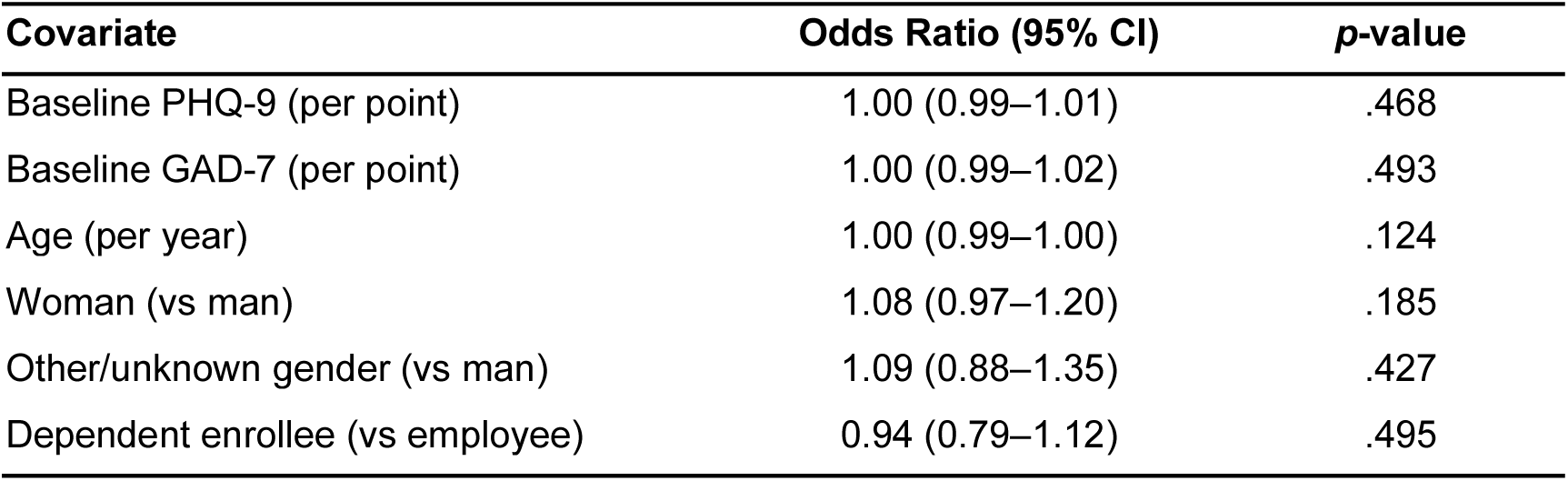
Predictors of any continuous care feature use among participants in intervention employers (n=8,021)

**Table S5.** Summary of Deviations from the Preregistered Analysis Plan.

| Preregistered Plan | Deviation | Rationale and Impact |
| --- | --- | --- |
| **Employer-level design:** Six employers assigned to the intervention condition and 12 matched employers assigned to control (2:1 matching). | **Expanded employer sample:** Twenty-five intervention employers matched to 75 comparison employers using a 1:3 matching ratio (total n=100 employers). | The scale of the real-world rollout exceeded initial projections, allowing inclusion of substantially more employers without altering the cluster-level assignment mechanism. Increasing the number of clusters improved statistical power, balance, and precision of employer-level estimates while preserving the original quasi-experimental design. |
| **Primary analytic focus on opt-in participants:** Primary analyses restricted to treatment participants who opted into session recordings and received the full AI feature set, with individual-level matching to controls. | **Intent-to-treat (ITT) primary analyses:** All participants in intervention employers were included regardless of feature opt-in, with exposure defined at the employer level. | Restricting analyses to opt-in participants would have introduced post-assignment selection bias due to partial uptake and consent-based feature access. The ITT approach preserves the original cluster-level assignment, avoids post-assignment selection bias, and yields more conservative estimates of program impact. Because ITT estimates average effects across all eligible participants, including those who did not engage with intervention features, estimated effects may be attenuated relative to treatment-on-the-treated contrasts; however, the substantially larger analytic sample increases statistical power to detect even modest but policy-relevant effects. This change strengthens causal interpretability relative to the preregistered plan. |
| **Clinical outcomes timing:** Assess changes in anxiety and depressive symptoms from baseline to a fixed 7-week post-treatment follow-up. | Clinical outcomes were modeled as continuous symptom trajectories up to 6 months following psychotherapy initiation, with follow-up restricted to observations occurring before the general availability launch date to avoid contamination. | A fixed 7-week endpoint would have discarded substantial valid follow-up data and reduced power. Modeling continuous trajectories better reflects real-world care patterns, improves estimation efficiency, and allows treatment effects to accumulate over clinically meaningful time horizons while preserving the original estimand of differential symptom improvement. |
| **Clinical improvement assessment:** Estimate change from baseline to a single post-treatment follow-up at 7 weeks, with treatment effects evaluated at that endpoint. | Clinical outcomes were analyzed as longitudinal symptom trajectories over time using group × time interactions. Robustness of estimated intervention effects were evaluated using large-scale falsification and difference-in-differences analyses applied to an independent pre-intervention cohort from the same employers in the year prior to launch. | Focusing inference on a single post-treatment endpoint would not have permitted direct tests distinguishing true intervention effects from secular trends or cohort differences. Modeling symptom trajectories enabled explicit tests of whether differential improvement emerged only following intervention rollout. Incorporating a full pre-launch cohort from the same employers allowed direct falsification of the treatment effect under identical model specifications, providing strong evidence that observed post-launch differences reflect the causal impact of the intervention rather than artifacts of timing, regression to the mean, or unobserved employer-level differences. This approach strengthens causal inference relative to the preregistered endpoint-based analysis. |
| **Small-sample cluster corrections** were planned due to a limited number of employer clusters (n=18). | Large-cluster mixed-effects models supplemented with falsification and difference-in-differences sensitivity analyses. | The final sample included a substantially larger number of clusters than anticipated (i.e., 100 employers), reducing reliance on small-sample corrections. The use of falsification tests and DiD models provides stronger evidence against residual confounding than cluster-adjustment alone, improving inferential robustness. |

### Deviations from Preregistration

This study was preregistered prior to the launch of the AI-enabled continuous care features, with the goal of prospectively specifying key design and analytic decisions (https://aspredicted.org/s7vr-6m7k.pdf). As is common in evaluations embedded within real-world program rollouts, several aspects of implementation evolved in ways that could not be fully anticipated at the time of preregistration. In particular, program development priorities, staggered employer launches, variation in provider and patient opt-in to session recordings and specific features, and the transition to general availability necessitated targeted analytic refinements to preserve internal validity, statistical power, and interpretability of treatment effects. Importantly, the core study design, comparison strategy, outcome measures, and causal estimands remained closely aligned with the preregistered plan. Where deviations occurred, they reflected practical constraints inherent to naturalistic implementation and were made to strengthen inference, reduce bias, or avoid underpowered or selectively defined analytic samples. All substantive deviations from the preregistered analysis plan are transparently documented below, along with their rationale and implications for interpretation.

---

**Figure S1.**
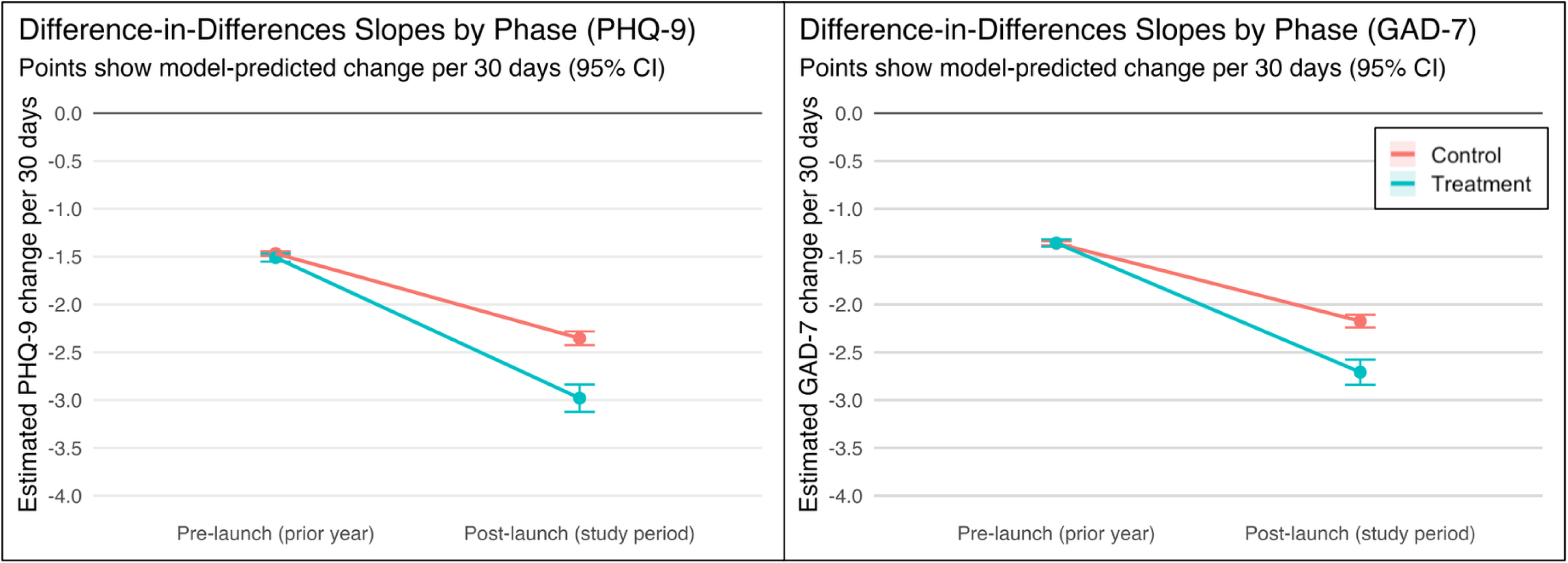
Difference-in-differences estimates of symptom improvement slopes. Slopes are shown for intervention and comparison groups in the pre-launch (prior year) and post-launch (current intervention period) phases. The difference-in-differences estimand corresponds to the differential change in improvement slopes from the pre- to post-launch period between groups.

## References

1. Schleider JL, Dobias ML, Mullarkey MC, Ollendick T. Retiring, Rethinking, and Reconstructing the Norm of Once-Weekly Psychotherapy. Adm Policy Ment Health Ment Health Serv Res. 2021;48(1):4–8. doi:10.1007/s10488-020-01090-7

2. Tiemens B, Kloos M, Spijker J, Ingenhoven T, Kampman M, Hendriks GJ. Lower versus higher frequency of sessions in starting outpatient mental health care and the risk of a chronic course; a naturalistic cohort study. BMC Psychiatry. 2019;19(1):228. doi:10.1186/s12888-019-2214-4

3. Ernst M, Hüwe L, Andreas S. Out of sight, out of mind? Intersession experiences as protective and risk indicators of suicidal ideation in the context of outpatient treatment. Psychother Res. Published online February 8, 2026:1–15. doi:10.1080/10503307.2026.2623939

4. Fernandez E, Salem D, Swift JK, Ramtahal N. Meta-analysis of dropout from cognitive behavioral therapy: Magnitude, timing, and moderators. J Consult Clin Psychol. 2015;83(6):1108–1122. doi:10.1037/ccp0000044

5. Swift JK, Greenberg RP, Tompkins KA, Parkin SR. Treatment refusal and premature termination in psychotherapy, pharmacotherapy, and their combination: A meta-analysis of head-to-head comparisons. Psychotherapy. 2017;54(1):47–57. doi:10.1037/pst0000104

6. Erbe D, Eichert HC, Riper H, Ebert DD. Blending Face-to-Face and Internet-Based Interventions for the Treatment of Mental Disorders in Adults: Systematic Review. J Med Internet Res. 2017;19(9):e6588. doi:10.2196/jmir.6588

7. Andersson G, Cuijpers P, Carlbring P, Riper H, Hedman E. Guided Internet-based vs. face-to-face cognitive behavior therapy for psychiatric and somatic disorders: a systematic review and meta-analysis. World Psychiatry. 2014;13(3):288–295. doi:10.1002/wps.20151

8. Thase Michael E., Wright Jesse H., Eells Tracy D., et al. Improving the Efficiency of Psychotherapy for Depression: Computer-Assisted Versus Standard CBT. Am J Psychiatry. 2018;175:242–250. doi:10.1176/appi.ajp.2017.17010089

9. Mohr DC, Lattie EG, Tomasino KN, et al. A randomized noninferiority trial evaluating remotely-delivered stepped care for depression using internet cognitive behavioral therapy (CBT) and telephone CBT. Behav Res Ther. 2019;123:103485. doi:10.1016/j.brat.2019.103485

10. Lan J, Qin Z, Niu H, Wang J. The Intervention Effects of Blended Cognitive Behavioral Therapy on Patients with Depression: A Meta-analysis. Cogn Ther Res. Published online October 31, 2025. doi:10.1007/s10608-025-10678-y

11. Graham S, Depp C, Lee EE, et al. Artificial Intelligence for Mental Health and Mental Illnesses: An Overview. Curr Psychiatry Rep. 2019;21(11):116. doi:10.1007/s11920-019-1094-0

12. Ali M, Ali S, Abbas Q, Abbas Z, Lee SW. Artificial intelligence for mental health: A narrative review of applications, challenges, and future directions in digital health. Digit Health. 2025;11:20552076251395548. doi:10.1177/20552076251395548

13. D’Alfonso S. AI in mental health. Curr Opin Psychol. 2020;36:112–117. doi:10.1016/j.copsyc.2020.04.005

14. Shimada K. The Role of Artificial Intelligence in Mental Health: A Review. Sci Insights. 2023;43(5):1119–1127. doi:10.15354/si.23.re820

15. Seidman AJ, Lannin DG, Heath PJ, Vogel DL. Setting the stage: The effect of affirming personal values before psychotherapy intake screenings on perceptions of self-stigma and self-disclosure. Stigma Health. 2019;4(3):256–259. doi:10.1037/sah0000140

16. Perlis RH. Artificial Intelligence and the Potential Transformation of Mental Health. JAMA Psychiatry. Published online January 14, 2026. doi:10.1001/jamapsychiatry.2025.4116

17. Li H, Zhang R, Lee YC, Kraut RE, Mohr DC. Systematic review and meta-analysis of AI-based conversational agents for promoting mental health and well-being. Npj Digit Med. 2023;6(1):236. doi:10.1038/s41746-023-00979-5

18. Ward EJ, Fragala MS, Birse CE, et al. Assessing the impact of a comprehensive mental health program on frontline health service workers. PLoS ONE. 2023;18(11 November):1–15. doi:10.1371/journal.pone.0294414

19. Bondar J, Babich Morrow C, Gueorguieva R, et al. Clinical and financial outcomes associated with a workplace mental health program before and during the COVID-19 pandemic. JAMA Netw Open. 2022;5(6):e2216349. doi:10.1001/jamanetworkopen.2022.16349

20. Hawrilenko M, Smolka C, Ward E, et al. Return on Investment of Enhanced Behavioral Health Services. JAMA Netw Open. 2025;8(2):1–14. doi:10.1001/jamanetworkopen.2024.57834

21. Graupensperger S, Hawrilenko M, Brown M, Baum G, Ward EJ, Chekroud A. Crisis Outreach, Treatment Engagement, and Outcomes After Suicide Risk Screening in a Comprehensive Mental Health Platform. Psychiatr Serv. Published online November 14, 2025. Accessed November 14, 2025. https://psychiatryonline.org/doi/10.1176/appi.ps.20250319

22. Ward, E. J., Hawrilenko, M., Wu, M., Novak, J., Brown, Millard, Chekroud, Adam. Large-Scale Evaluation of Provider-Patient Matching in an Employer-Sponsored Mental Health Program. Published online In Revision.

23. Baum, Graham, Hawrilenko, Matt, Cascalheira, Cory, et al. Mental Health Service Use and Equity in a Comprehensive Employer-Sponsored Benefit Program: A Retrospective Cohort Study. Popul Health Manag. Published online 2026. doi:10.1177/19427891261420041

24. Kroenke K, Spitzer RL, Williams JBW. The PHQ-9: Validity of a brief depression severity scale. J Gen Intern Med. 2001;16:606–613.

25. Spitzer RL, Kroenke K, Williams JBW, Löwe B. A brief measure for assessing generalized anxiety disorder: The GAD-7. Arch Intern Med. 2006;166:1092–1097. doi:10.1001/archinte.166.10.1092

26. McMillan D, Gilbody S, Richards D. Defining successful treatment outcome in depression using the PHQ-9: A comparison of methods. J Affect Disord. 2010;127(1-3):122–129. doi:10.1016/j.jad.2010.04.030

27. Toussaint A, Hüsing P, Gumz A, et al. Sensitivity to change and minimal clinically important difference of the 7-item Generalized Anxiety Disorder Questionnaire (GAD-7). J Affect Disord. 2020;265:395–401. doi:10.1016/j.jad.2020.01.032

28. Constantino MJ, Muir HJ, Gaines AN, Ouimette K. Hope and expectancy factors. In: The Field Guide to Better Results: Evidence-Based Exercises to Improve Therapeutic Effectiveness. American Psychological Association; 2023:131–153. doi:10.1037/0000358-007

29. Rozenkrantz L, Laskorunskyi O, Zilcha-Mano S, Dattner I. Dynamic expectancies: The independent role of within-person change in outcome expectancy in predicting overall treatment outcomes in psychotherapy for depression. Psychother Res. 2025;0(0):1–11. doi:10.1080/10503307.2025.2519574

30. Elliott R, Bohart AC, Watson JC, Murphy D. Therapist empathy and client outcome: An updated meta-analysis. Psychotherapy. 2018;55(4):399–410. doi:10.1037/pst0000175

31. Kazantzis N, Whittington C, Zelencich L, Kyrios M, Norton P, Hofmann S. Quantity and Quality of Homework Compliance: A Meta-Analysis of Relations With Outcome in Cognitive Behavior Therapy. Behav Ther. 2016;47. doi:10.1016/j.beth.2016.05.002

32. Dong L, Zhao X, Ong SL, Harvey AG. Patient recall of specific cognitive therapy contents predicts adherence and outcome in adults with major depressive disorder. Behav Res Ther. 2017;97:189–199. doi:10.1016/j.brat.2017.08.006

33. Ryum T, Bennion M, Kazantzis N. Homework as a driver of change in psychotherapy. J Clin Psychol. 2024;80(4):733–743. doi:10.1002/jclp.23627

34. Cuijpers P, Harrer M, Miguel C, Karyotaki E, Papola D. Does baseline severity interact with the effects of psychotherapy for depression? A meta-analytic review. J Affect Disord. 2026;399:121106. doi:10.1016/j.jad.2025.121106

35. Bruijniks S, Harvey AG, Hollon SD, et al. Use and Effects of Therapist Memory Support Strategies in Cognitive Behavioral Therapy and Interpersonal Psychotherapy for Depression. Cogn Ther Res. 2025;49(5):1017–1030. doi:10.1007/s10608-024-10569-8

36. Harvey AG. Maximizing benefits from evidence-based psychological treatments: Memory support and habit formation as key strategies. Behav Res Ther. 2025;191:104767. doi:10.1016/j.brat.2025.104767

